# A Re-Appraisal of Three Network Meta-Analyses to Explain the Discrepancy in Findings for the Efficacy of Fluoxetine for the Treatment of Depression in Children and Adolescents

**DOI:** 10.1101/2025.09.07.25334757

**Authors:** Richard Lyus, Florian Naudet, Gert van Valkenhoef, Martin Plöderl

## Abstract

**Objective:** To explain discrepant findings for fluoxetine’s efficacy in three influential network meta-analyses (NMAs) of treatments for pediatric depression, which led to conflicting clinical recommendations.

**Design:** Critical appraisal and re-analysis of three published NMAs.

**Data sources:** NMAs published in two Lancet journals and in Cochrane, together with the trial datasets reported therein.

**Data synthesis:** We compared efficacy estimates for fluoxetine versus placebo across NMAs. We identified and assessed an outlying trial included only in the Lancet NMAs using the INSPECT-SR instrument, and re-analysed the NMAs with and without this trial.

**Results:** The larger effects reported in the Lancet NMAs (SMD -0.51, 95% CrI -0.99 to -0.03; and -0.51, -0.84 to -0.18) were driven by an inconsistent fluoxetine–placebo–nortriptyline loop, which the original NMA authors could not explain. We identified the cause of the inconsistency as a small outlier trial of fluoxetine versus nortriptyline that reported an implausibly large effect size (SMD > 4) favouring fluoxetine, and which was not included in the Cochrane NMA. Excluding this trial from the Lancet NMA datasets resolved the inconsistency and yielded efficacy estimates for fluoxetine that closely matched the Cochrane NMA (SMD -0.20, 95%CI -0.28 to -0.11). The outlier trial also showed multiple methodological concerns suggesting low trustworthiness.

**Conclusion:** Discrepancies between the three NMAs were explained by the indirect influence of a single small trial with extreme and unreliable results. Removing this trial reconciled the Lancet NMAs with the Cochrane NMA, yielding a more reliable estimate of fluoxetine efficacy versus placebo. It also resolved the inconsistency. This case illustrates how inclusion of a single small problematic trial can substantially distort the clinically important results of NMAs. Our findings may alter the clinical risk/benefit assessment of fluoxetine for this indication.

**Other**: No specific funding was involved in the study.

**KEY MESSAGES:** *What is already known on this topic:*

● Three network meta-analyses frequently inform clinical guideline recommendations for the use of fluoxetine to treat depression in children and adolescents.
● However, these studies have discrepant findings for fluoxetine versus placebo, with the two earlier NMAs reporting an SMD approximately -0.5, and the other reporting a mean difference equivalent to an SMD of approximately -0.2.
● Such discrepancies require investigation because network meta-analyses should be reproducible syntheses of the available evidence and different results may have different clinical implications.
● Outlying or problematic trials in meta-analyses can distort results.

*What this study adds:*

● We identified that the cause of the discrepant findings was the indirect influence of a single small study of fluoxetine versus nortriptyline that reported an unusually large effect size favouring fluoxetine. This study was not included in the more recent NMA, which excluded studies of tricyclic antidepressants. In addition to the extreme effect size, this study has other concerning features that call into question its credibility.
● Our findings demonstrate how significantly a single small study with outlying results can impact the clinically important findings of NMAs. While it is known that meta-analyses can be affected by the inclusion of retracted studies, less is known about the impact of single unretracted studies with extreme findings and other concerning features.
● Our experience demonstrates that researchers seeking to critically appraise studies with concerning features, and the evidence syntheses that include them, may face barriers when raising their concerns with study authors and journal editors.

*How this study might affect research, practice, or policy:*

● Our findings may alter the clinical risk/benefit assessment of fluoxetine for this indication.
● Our findings show how evidence from small studies with outlying results can influence the most clinically important findings of NMAs, and highlight the need for NMA methodology to include assessment for outliers and for the credibility of individual trial results.

## INTRODUCTION

Treatment recommendations rely on high-quality and trustworthy evidence and its synthesis in systematic reviews and meta-analyses. Discrepant findings in evidence syntheses require explanation, particularly if the findings have differing clinical implications. It seems that this problem applies to fluoxetine for the treatment of pediatric depression.

Depression affects a significant proportion of children and adolescents and a recent review estimated the global point prevalence at 8%.[1] Worldwide, pharmacotherapy for this indication is initiated by a range of clinicians, including Family Physicians, Paediatricians, Psychiatrists, and nonmedical prescribers.[2] Antidepressant prescription rates in children and adolescents are ever increasing and fluoxetine is among the most frequently prescribed antidepressants, [3–6] likely because fluoxetine is recommended as the preferred medication in major guidelines.[7]

Major guidelines and similar publications[7–10] base their recommendations for fluoxetine as first-line medication on two network meta-analyses (NMAs) published in Lancet group journals by Cipriani et al. (2016)[11] and Zhou et al. (2020)[12] (Henceforth simply referred to as Ciprani and Zhou). Cipriani reported a standardised mean difference (SMD) for depression symptom reduction for fluoxetine versus placebo of -0.51 (95% Credible Interval [CrI] -0.99 to -0.03). Zhou reported a nearly identical SMD -0.51 (95% CrI -0.84 to -0.18). The confidence in the efficacy findings for fluoxetine was rated as ‘very low’ in both NMAs.

In contrast, the more recent NMA by Hetrick et al. (2021) (henceforth simply Hetrick) and published by Cochrane, produced a different result.[13] Efficacy results for depression symptom reduction were restricted to trials using the most common symptom rating scale, the Children’s Depression Rating Scale-Revised (CDRS-R, range 17 to 113 points), and Hetrick did not report an SMD. Instead they reported a mean difference (MD) of -2.8 points on the CDRS-R (95% Confidence Interval [CI] -4.12 to -1.56); but using a standard deviation (SD) of 14.471 as suggested by Hetrick, the SMD can be calculated as -0.20 (−0.28 to -0.11). The confidence in the finding was rated as ‘moderate’.

While the confidence intervals between the Lancet and Cochrane NMAs overlap and statistical significance was found in all three NMAs, the point-estimate and the lower end of the confidence interval were reduced by approximately half in the Cochrane NMA. Moreover, fluoxetine’s efficacy was now clinically equivalent with placebo (even when considering the whole range of the confidence interval) according to the predetermined criteria used by Hetrick, i.e. +/- 5 CDRS-R points (approximately equivalent to SMD 0.35). The appropriate range for equivalence can be debated, but Hetrick’s threshold is at the the smaller end of any such range, both by Hetrick’s own criteria (they suggested a range at least 50% of 10-15 points as a minimum) and by common estimates of the minimal important difference in antidepressant research.[14] Consequently, recommendations for fluoxetine as a first line treatment are at odds with the findings of the Cochrane NMA. In fact, the findings arguably support the view that fluoxetine should not be used at all, an argument that the authors acknowledged in their conclusion but refrained from endorsing themselves.

To our best knowledge, these discrepancies have not been acknowledged or explained in the scientific literature, clinical guidelines, or the Cochrane NMA itself, despite some overlap of authorship in the three NMAs.

Furthermore, from a methodological point of view, discrepant findings between NMAs assessing the same treatments require explanation, because NMAs are supposed to be reproducible evidence syntheses. Potential reasons for differences between NMAs include varying inclusion criteria or statistical methods, and errors. For example we have frequently identified what we call the ‘standard error’, that is, when the standard error is conflated with the standard deviation, leading to erroneously large effect sizes. Another explanation of the discrepancy would be the inclusion in the more recent NMA of more recent studies with smaller effect estimates. Furthermore, more recent trials often have multiple arms, with a novel drug, placebo, and fluoxetine as an established comparator drug. This reduces the probability of receiving placebo and may change expectancy effects and unblinding bias.[7] However, when we investigated this in detail, the effect of more recent trials that were included in the the more recent Cochrane NMA was not sufficient to explain the discrepant findings in the three NMAs.[7] Therefore, we aimed to further explore why the fluoxetine-placebo effect sizes differed in the Lancet NMAs and Hetrick by critically examining and re-analyzing the NMAs.

## METHODS

### Data collection

RL, FN, and MP examined the NMAs and sought associated data and code. The complete dataset for Cipriani is freely available through the <u>Oxford University Research Archive</u>. The dataset for Zhou is freely available through <u>Mendeley</u>. For Hetrick, the full dataset was not publicly available. We requested this from the lead author and from Cochrane, but the full dataset had not been retained and only a partial dataset was provided (Sarah Hetrick and Nick Meader, personal communication, 27 May 2025). MP and RL compiled a dataset of the studies that were included in Hetrick, combining the partial data provided by the authors with additional data that we extracted from the Zhou dataset (provided online for details), and following the preferences for data sources and types that were stated by Hetrick.

### Critical Analysis of an Outlier Trial

When a trial with an extreme effect size and various other features that undermined its credibility was identified among the datasets of the NMAs, we formally assessed it using outlier detection tests and the INSPECT-SR (INveStigating ProblEmatic Clinical Trials in Systematic Reviews)) framework.[15] We contacted the authors of the study, the journal editor, and the publisher, to raise our concerns.

### Critical analysis of the Three NMAs

We adopted a constructivist approach, engaging in a critical analysis of the above materials, followed by a collaborative discussion among three of the authors (RL, MP, FN). We structured our analysis around NMA methodology, network geometry, and inconsistency.[16]

### Re-analysis of the Three NMAs

#### Effect measures

We re-analysed efficacy estimates for fluoxetine as assessed by change in depression symptoms with placebo as reference group, because it was for this outcome that we observed discrepancies across the three NMAs. To facilitate comparison, we used SMDs in the synthesis and presentation of results.

#### Synthesis methods

One of the authors (MP) undertook a full reanalysis of the original Cipriani dataset, including sensitivity analysis to control for the effect of a single trial with outlying results - see below. When discrepancies persisted between the original NMA results and our reproduced results using the full dataset, MP involved an expert in Bayesian statistics NMA (GvV). We then conducted a re-analysis of all three NMAs.

The dataset from Cipriani provided the relevant information (mean change from baseline, SD, sample size) for all arms and trials and was thus ready to use for the re-analysis. The dataset from Zhou included similar information; however, there was variation in the assignment of treatment arms to results without supplementary information to explain the assignments. Therefore, we inspected each trial in the dataset to ensure matching of treatment arms to results. For re-analysis of Hetrick we created a dataset as described above. In trials with multiple doses we combined arms following the formula provided in the Cochrane Handbook.[17] There was no missing data. Estimates of direct and indirect summary measures were compiled and presented through tabulation.

#### Analyses

For all re-analyses, we initially aimed to achieve computational reproducibility by using the same data and code as in the published articles. We twice contacted the corresponding authors of all three studies to request the full code used in their analyses (Cipriani and Zhou published only part of their code; Hetrick published none). We did not receive any code. Therefore, we wrote an analytical code, which is available via the Open Science Framework (<u>OSF</u>).

We conducted Bayesian NMAs using a random effects model for the Lancet studies’ datasets, using all available information including details of priors to emulate their analyses (vague priors with a uniform distribution between 0 and 1 for the heterogeneity SD, and a normal with mean 0 and SD = 100 for all other prior distributions). We used R version 4.3.0[18] with R’s “gemtc” package version 1.1.[19] Model convergence was assessed with visual inspection of trace-plots and Gelman plots. We could not achieve perfect computational reproducibility so we conducted a sensitivity analysis using a frequentist approach to assess the robustness of the Bayesian results for Cipriani and Zhou.

We conducted a frequentist random effects NMA for the Hetrick dataset, as in the original publication, using R’s “netmeta” package version 3.2-0.[20]

To assess the impact of a single study with outlying results on efficacy estimates, heterogeneity, and consistency, we re-evaluated the two Bayesian NMAs with and without this study. We also ran tests of outliers by investigating standardized residuals and Mahalanobis distance with R’s NMAoutlier package[21] for the Cipriani and Zhou data.

### Open Science and reporting

This exploratory study did not have a pre-registered protocol. All data and code are either publicly available (links in text) or on the <u>OSF</u>. While there is no specific reporting guideline for this type of analysis, we adapted the PRISMA-NMA framework for reporting findings.[16]

### Patient Involvement

No patients were involved in this study. All results and their potential implications will be discussed on 9th September 2025 by the RestoRes (Research Integrity in Biomedical Research) project’s external advisory committee, which includes various stakeholders such as patient representatives, editors, and journalists. The committee will also address the most appropriate ways to disseminate these results to the public following their publication.

## RESULTS

### Critical analysis of the Three NMAs

#### NMA methodology

Table 1 shows the key characteristics of the three NMAs regarding their fluoxetine-placebo efficacy findings. Cipriani and Zhou used a Bayesian approach while Hetrick used a frequentist one. This would not be expected to account for substantial differences in a single finding, particularly when the Bayesian analyses used vague priors.

**Table 1.** Characteristics of three network meta-analyses of medication for treatment of depression in children and adolescents focusing on fluoxetine versus placebo efficacy findings for symptom reduction. SMD is for change in depression rating scales. *Hetrick et al. reported only mean difference (MD) and did not report an SMD; the SMD here is calculated using a standard deviation (SD) 14.471, as the authors suggest based on their previous version of the NMA, Hetrick et al. (2012).

|  | **Cipriani et al. (2016)** | **Zhou et al. (2020)** | **Hetrick et al. (2021)** |
| --- | --- | --- | --- |
| **Interventions considered** | Antidepressants | Antidepressants and psychotherapy | New generation antidepressants |
| **Efficacy Outcomes Included for Depression Symptoms** | CDRS-S, CDI, BDI | 'Standardised depressive symptom scales.' | CDRS-R only |
| **Total # Studies** | 34 | 71 | 26 |
| **# Studies including a fluoxetine arm** | 10 | 14 | 9 (8 used the CDRS) |
| **# Studies including fluoxetine and placebo arms** | 8 | 9 | Not stated; we calculated 9 |
| **Flx v Pbo network SMD** | -0.51 (95% CrI -0.99 to -0.03) | -0.51 (95% CrI -0.84 to -0.18) | -0.20* (95% CI -0.28 to -0.11) using -2.84 points on CDRS-R and SD = 14.471 |
| **Pre-defined equivalence range** | None | None | +/-5 points on CDRS-R |
| **Provided direct and indirect estimates** | In appendix | In appendix | No |
| **Confidence Rating for Flx v Pbo SMD** | Very low | Very low | Moderate |

We considered whether differences in study populations would explain the inconsistent findings, particularly the possibility that Hetrick included more recent studies with direct comparisons of fluoxetine versus placebo that were not available for the earlier NMAs. Two such studies (Findling et al. 2020, 2022) were included only in Hetrick, both of which used fluoxetine as a control arm alongside placebo and an investigational drug (vilazodone and vortioxetine respectively). Findling (2020) reported an MD for fluoxetine versus placebo of -2.3 points (p = 0.14) on the CDRS-R, while Findling (2022) reported an MD of -3.73 points (p = .015) favouring fluoxetine.[22,23]. However, in our pairwise meta-analytic aggregation of fluoxetine trials over time, published elsewhere,[7] these two studies had no notable effect on the point estimate.

#### Network geometry

We did not identify any major differences in the network geometry for medication. Zhou included studies of psychotherapy as well as medication. In all three NMAs, placebo was the most common comparator for drug trials and fluoxetine the most studied drug.

#### Inconsistency

Both Lancet NMAs reported inconsistency between the direct and indirect results for fluoxetine versus placebo, identified using the node-splitting method (separating direct and indirect estimates for each comparison), i.e. the direct and indirect estimates were statistically significantly different. The supplementary material for both NMAs showed that there were very large differences between the direct and indirect results for the fluoxetine-notriptyline-placebo loop (summarised in Table 2). The effect sizes for the direct comparison between nortriptyline versus fluoxetine, and for the indirect comparison between nortriptyline versus placebo, were very large (SMD ≈ 4).

**Table 2.** Evaluation of the inconsistency by node-splitting model in Cipriani et al. and Zhou et al. for the comparisons comprising the fluoxetine-nortriptyline-placebo loop for efficacy on depression symptoms.

|  |  | Direct |  | Indirect |  | Difference |  |  |
| --- | --- | --- | --- | --- | --- | --- | --- | --- |
| Study | Comparison | SMD | SE | SMD | SE | SMD | SE | p |
| Cipriani 2016 | PBO-FLX | 0.26 | 0.14 | 1.40 | 0.48 | -1.14 | 0.50 | 0.0209 |
|  | PBO-NOR | 0.11 | 0.25 | -3.96 | 0.62 | 4.07 | 0.67 | <0.001 |
|  | NOR-FLX | 4.22 | 0.62 | 0.15 | 0.26 | 4.07 | 0.67 | <0.0001 |
| Zhou 2020 | PBO-FLX | 0.24 | 0.12 | 1.26 | 0.30 | -1.02 | 0.33 | 0.0019 |
|  | PBO-NOR | 0.12 | 0.29 | -3.93 | 0.65 | 4.06 | 0.71 | 0.0000 |
|  | NOR-FLX | 4.22 | 0.64 | 0.17 | 0.30 | 4.06 | 0.71 | 0.0000 |

The inconsistent results included a very large direct result for the comparison of fluoxetine versus nortriptyline, SMD 4.22 in both Lancet NMAs, and we noted that effects of this size are unheard of in studies of antidepressants. The largest effect size reported in a 2018 NMA of medication for treatment of depression in adults, including drug-drug comparisons, was amitriptyline versus placebo, with SMD (pooled) -0.48,[24] and in a review of various psychiatric drug treatments the maximum reported effect size was SMD 1.12.[25] The pattern of inconsistency in the fluoxetine-nortriptyline-placebo loop indicated that the efficacy result for fluoxetine versus placebo had been indirectly informed by this very large SMD for fluoxetine versus nortriptyline.

#### Identifying the Source of the Large Effect Size in The Lancet NMAs

To identify the source of the large SMD for the direct fluoxetine versus nortriptyline comparison in the Lancet NMAs we reviewed the included studies to find head-to-head trials of these drugs. In both NMAs there was only one such study, Attari et al. (2006).[26] This was a small trial of 40 patients randomized to fluoxetine (n=20) or nortriptyline (n=20) without a placebo arm, undertaken at a single site in Iran. The study used the self-rated Children’s Depression Inventory (CDI; range 0-54) to measure change in depression symptoms. It reported a mean change score of -10.9 (SD 2.6) for fluoxetine and -2.6 (SD 0.8) for nortriptyline, giving an MD on the CDI of 8.35 points favoring fluoxetine, corresponding to SMD -4.24 (95% CI -5.37 to -3.11, see <u>OSF</u>).

To further scrutinize this large effect size we considered Hetrick’s subgroup analysis of three studies that used the CDI. The raw mean difference for drug versus placebo was -1.30 points (95% CI -5.87 to 3.27) for fluoxetine, -0.43 (−2.91 to 2.05) for paroxetine, and -0.28 (−3.72 to 3.16) for citalopram. The difference of 8.35 points reported by Attari et al. for fluoxetine versus nortriptyline is obviously much greater. We then considered the Attari et al. effect size in the context of the other effect sizes in the Lancet NMA datasets (for all rating scales). For the Cipriani dataset, Attari’s effect size of -4.24 is more than 4 standard deviations from the pooled mean effects and more than four times the next largest effect size of 0.9 (Figure 1). Attari’s outlier status was further confirmed by Mahalanobis plot and standardised residuals plot (Figures S2, S3).

**Figure 1.**
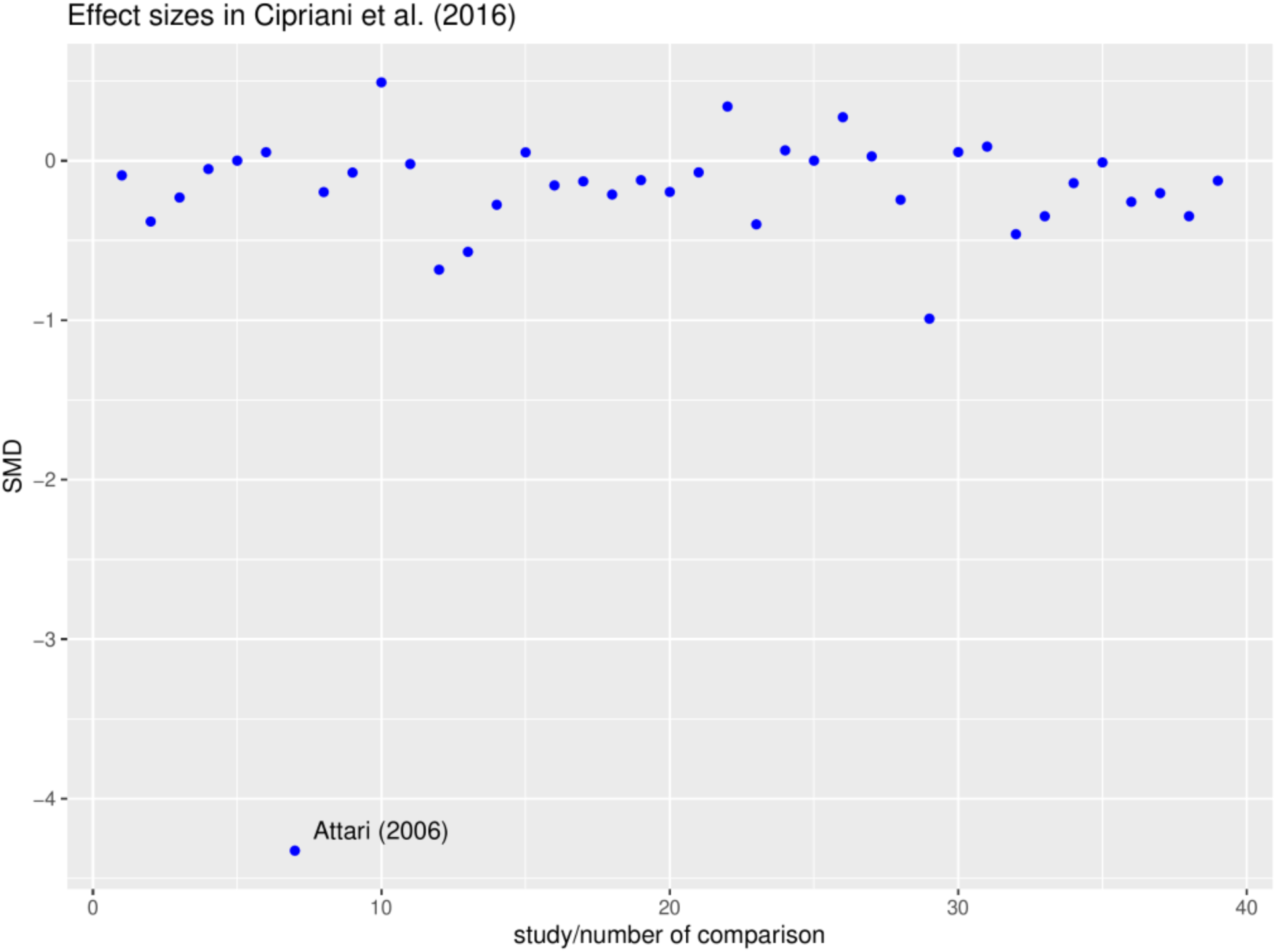
Effect sizes in Zhou et al. (2020)

We examined the supplementary material of the Lancet NMAs to see if attempts were made to identify outlying studies. The only assessment we could identify were funnel plots, which are primarily used to identify publication bias but may secondarily identify outliers. However, in both Lancet NMAs the effect sizes had been centred on the pooled *comparison-specific* effects, not on the pooled effect for the full dataset, so the comparison for nortriptyline versus fluoxetine was plotted at zero because Attari et al. was the only study for this comparison (i.e. it was compared to itself).

Next, the possibility was considered that the findings of Attari et al. could be explained by nortriptyline being genuinely (and greatly) inferior to placebo, but this was not supported by pairwise comparisons of nortriptyline versus placebo in the Lancet NMAs, both of which reported SMD -0·11 (95%CrI -0·55 to 0·34). This result was informed by two trials of nortriptyline versus placebo (Geller et al. 1990, n=31; Geller et al. 1992, n=60).

#### Critically Assessing the Outlier Trial

Considering the extremity of the effect size reported by Attari et al., we proceeded to analyse this study for other concerning features that might bring into question the credibility of its findings. To guide this analysis we used the preliminary version of INSPECT-SR. We identified concerning features in two of four domains, particularly regarding contradictions in the data (see Table 3). The statistical results reported in the study were incompatible with each other, i.e. impossible. The extreme effect size based on change scores was more than four times larger than the effect size based on endpoint scores (SMD = -0.93 [95% CI -1.59 to -0.28], see <u>OSF</u>), and also differed from effect-sizes calculated from baseline and endpoint values and assuming a range of correlations. To produce the reported effect size the correlation between pre-post scores would have to be almost perfect (r > 0.98; see figure S1 in Appendix and <u>OSF</u>) and this is not plausible. Furthermore, we noted unusually small SDs for the change scores and larger SDs for the groups based on pre- and post-scores. These features call into question the trustworthiness of the data in Attari et al.

**Table 3.** INSPECT-SR Checklist for the trial by Attari et al. [26]

| Check | Response | Comments |
| --- | --- | --- |
| **Domain: Inspecting post-publication notices**<br>*Overall domain judgement: No concerns* |  |  |
| Does the study have an associated retraction? | No |  |
| Does the study have an associated expression of concern or other relevant post publication notice? | No | The study is not listed in PubMed and has no Digital Object Identifier, thus it is not possible to raise concern via platforms such as PubPeer. |
| Do other studies by the research team highlight causes for concern (associated retractions, expressions of concern, relevant post-publication notices?) | No |  |
| **Domain: Inspecting conduct, governance and transparency**<br>*Overall domain judgement: Some concerns* |  |  |
| Are there concerns relating to ethical approval? | Yes | Ethical approval not mentioned. |
| Are there concerns relating to the timing or absence of study registration? | Unclear | No registration but study was published 2006. |
| Are there important inconsistencies between the publication and the registration documents? | N/A | No protocol is available. |
| Is the recruitment of participants implausible? | No | 40 patients in two years at a single centre. |
| Are the reported methods implausible considering the reported resources? | Unclear | No information on the resources. |
| **Inspecting text and figures** |  |  |
| *Overall domain judgement: No concerns* |  |  |
| Is there any plagiarised text, or text that is incompatible with the study? | No |  |
| Is there evidence of manipulation or duplication of figures? | No |  |
| **Inspecting results in the study** |  |  |
| *Overall domain judgement: Serious concerns* |  |  |
| Are there any unexplained discrepancies between reported data and participant inclusion criteria? | No |  |
| Are numbers of participants allocated to each group implausible given the allocation method? | No |  |
| Are any baseline data implausible? | No |  |
| Are there any discrepancies between results reported in figures, tables, and text? | Yes | Minimal differences between text and data in Table 1: Text states no difference between groups for ethnicity; ethnicity is not reported in the table. |
| Are the numbers of participants lost to follow-up implausible? | No |  |
| Are there any unexplained inconsistencies in the numbers of participants? | Yes | 'The dropout in nortriptyline group and fluoxetine group were 4 and 2 respectively, during the study. So, they were replaced by new cases.' Replacement of dropouts is rarely done in RCTs, and no information is given if this was pre-specified or done post-hoc. Moreover, it is not clear if replacements were randomized. |
| Are any outcome data, including estimated treatment effects, implausible? | Yes | SMD 4.3 based on change scores is implausible and substantially deviates from the SMD 0.93 based on endpoint scores.<br>To produce the reported effect size the correlation between pre-post scores would have to be almost perfect ($r > 0.98$).<br>SD 0.8 for change score in fluoxetine group is implausible. |
|  |  | The small SD 0.8 for the fluoxetine group suggests that the response was very similar for all the patients in the group; this is unlikely to be compatible with the following: 'At the endpoint (8th week), 10 cases (50%) didn't meet the criteria of Major Depression based on DSM-IV in fluoxetine group.'<br>SDs for pre-post groups and implausibly different from SDs for change scores. |
| Are the means and variances of integer data impossible? | No | All reported mean values were checked with the GRIMMER test. |
| Are there errors in statistical results? | Yes | The p-value of the t-test for the main result is given as $p = 0.004$, but it should be 0.0052, and this cannot be explained with rounding issues.<br>SDs for pre-post scores are unlikely to be compatible with SD scores for change scores. |
| Are any other contradictions implied by the data? | No | No |
| Are there inconsistencies in descriptions of methods and results across publications describing the study? | Yes | Unexplained replacement of dropouts. |
| **Overall study judgement: Serious concerns** |  |  |

However, even if concerns about trustworthiness are set aside, Attari et al. is obviously an outlier. The Cochrane Handbook recommends that, ‘It is advisable to perform analyses both with and without outlying studies as part of a sensitivity analysis’.[17] No such sensitivity analysis was undertaken in either of the Lancet NMAs. We noted that because Hetrick excluded studies of tricyclics from their analysis, Attari et al. would not have been included. We therefore hypothesised that this study might explain the discrepant findings for the efficacy of fluoxetine between the Lancet NMAs and Hetrick, and proceeded to undertake a sensitivity analysis of the Lancet NMA datasets with and without this study.

### Re-analysis of the Three NMAs With and Without the Outlier Trial

We re-analysed the Cipriani and Zhou datasets with and without Attari et al. using a Bayesian approach, to see how this study affected the fluoxetine efficacy estimates. Table 4 shows the results of the original analyses and all re-analyses with and without Attari et al. We were able to closely reproduce the original findings of Cipriani and Zhou, except for some indirect estimates that were larger in our analyses. We reproduced the statistically significant inconsistency between direct and indirect comparisons for fluoxetine for both NMAs.

**Table 4.** Results of the re-analysis of Cipriani et al. for fluoxetine efficacy versus placebo with and without Attari et al. 2006. Efficacy estimates are given as SMDs for Cipriani and Zhou and CDRS-points for Hetrick. Intervals in brackets are 95% credible intervals (CrI) in Bayesian analysis or 95% confidence intervals (CI) in frequentist analysis. CrIs from the ‘Original NMA’ were calculated using +/- 1.96 multiplied with the reported SE; the SMD of the original Hetrick et al. finding was calculated using the suggested SD by Hetrick (see text for details).]

| Study | Analysis | Direct Efficacy Estimate SMD (95% CrI/CI) | Indirect Efficacy Estimate SMD (95% CrI/CI) | p-value for the test of inconsistency | Combined Efficacy Estimate SMD (95% CrI/CI) |
| --- | --- | --- | --- | --- | --- |
| Cipriani 2016 | Original NMA | -0.26 (-0.54, 0.02) | -1.40 (-2.34 to -0.46) | 0.021 | -0.51 (-0.99 to -0.03) |
|  | Re-analysis with Attari | -0.24 (-0.68 to 0.20) | -2.1 (-3.3 to -1.1) | 0.00278 | -0.51 (-0.99 to -0.03) |
|  | Re-analysis without Attari | -0.27 (-0.46 to -0.07) | -0.14 (-0.58 to 0.86) | 0.72269 | -0.26 (-0.43 to -0.09) |
| Zhou 2020 | Original NMA | -0.24 (-0.48 to 0.00) | -1.26 (-1.85 to -0.67) | 0.002 | -0.51 (-0.84 to -0.18) |
|  | Re-analysis with Attari | -0.23 (-0.56 to 0.09) | -1.9 (-2.70 to -1.10) | 0.00014 | -0.49 (-0.82 to -0.16) |
|  | Re-analysis without Attari | -0.24 (-0.47 to -0.01) | -0.68 (-1.3 to 0.02) | 0.23341 | -0.29 (-0.50 to -0.07) |
| Hetrick 2021 | Original NMA (raw CDRS score) | Not reported | Not reported | Not reported | -2.80 (-4.12 to -1.56) |
|  | Re-analysis raw CDRS score | -3.04 (-4.47 to -1.61) | -4.68 (-12.17 to 2.82) | 0.6739 | -3.09 (-4.50 to -1.69) |
|  | Re-analysis SMD | -0.26 (-0.38 to -0.14) | -0.39 (-1.01 to 0.24) | 0.7009 | -0.27 (-0.38 to -0.15) |

Notably, exclusion of the trial by Attari et al. resolved the inconsistency for all three comparisons comprising the fluoxetine-nortriptyline-placebo loop. The network efficacy estimates for fluoxetine versus placebo were substantially smaller without Attari et al., with SMD -0.26 (−0.43 to -0.09) for Cipriani, and -0.29 (−0.50 to -0.07) for Zhou , i.e. much closer to the result of Hetrick et al., which we had calculated as -0.2 using the MD and suggested SD from the original NMA, and which was -0.27 (−0.38 to -0.15) in our re-analysis using our reconstructed Hetrick dataset. For the frequentist approach we also found comparable results in the sensitivity analysis for Cipriani and Zhou (Table S1).

In our re-analysis of Hetrick there was no inconsistency between direct and indirect comparisons for fluoxetine versus placebo (SMD -0.26 and -0.39, p = 0.70), in-keeping with the original NMA.

### Raising Concerns

We twice contacted the authors of Attari et al. to raise our concerns about the validity of their data but received no response. We then twice contacted the editor of the journal in which it was published, and received a response which said the editor had forwarded our concerns to the authors. We followed this up with further emails but at the time of writing we have received no response.

The journal in which Attari et al. was published has no Digital Object Identifier and is not listed in PubMed so we are unable to post commentary on, for example, PubPeer. We submitted our findings (as an earlier version of this article) to Lancet Psychiatry, who sent our manuscript for peer review and shared our findings with The Lancet and with The Lancet Group Research Integrity Team. Based on the reports of two reviewers, The Lancet Group journals rejected our manuscript and advised us that they felt no further action was required on their part and they considered the matter closed. They suggested that if we had concerns about Attari et al. we should raise these with the relevant authors or editors

## DISCUSSION

### Principal Findings

We have explained the discrepant findings between the Lancet and Cochrane NMAs: the inclusion in the former of Attari et al., which increased the overall estimate for fluoxetine versus placebo, through indirect comparisons. In our re-analysis we found that removing this trial resolved the discrepant findings for fluoxetine’s efficacy, and it resolved the inconsistency between direct and indirect estimates in the Lancet NMAs. An important finding from our analysis is that one small outlying trial can substantially bias the overall results in an NMA, including the most clinically impactful results.

The question arises of why Attari was not identified as a problematic trial in the original Lancet publications. While Attari et al. was rated as having a moderate risk of bias by Cipriani and an unclear risk of bias by Zhou, two important features of this trial remained undetected. First, the extreme effect size in Attari is clearly a statistical outlier. Second, it also has multiple concerning features which undermine its reliability so significantly that it may need to be corrected or retracted. The inconsistency it created was acknowledged as problematic in both NMAs, and they offered various hypotheses as explanation, but the cause was not identified and consequently sensitivity analysis was not done.

Our re-analysis supports Cochrane’s recommendation to run sensitivity analyses when there are statistical outliers, but shows that obvious outliers can remain unacknowledged even in meta-analyses by large teams of experienced researchers. Furthermore, our findings suggest that existing risk-of-bias analyses are not sufficient to detect problematic trails. Systematic reviews typically assess the methodological quality of the included trials, but do not consider whether the data themselves can be trusted. The importance of assessing the trustworthiness of findings, using a tool such as INSPECT-SR,[15] is increasingly recognised as an essential part of ensuring the integrity of research syntheses.[27]

Our re-analysis also has implications regarding the evaluation of fluoxetine’s efficacy. While the confidence or credible intervals in all three NMAs excluded the null-effect, the point-estimates and lower margin of the credible interval in the Lancet NMAs were reduced by about 50% from the original results, in line with the findings by Hetrick. This efficacy estimate puts fluoxetine much more clearly in the the range of clinical equivalence with placebo in all three NMAs, using both the Hetrick definition and using established estimations in antidepressant research.[14]

Our experience of sharing our findings with authors and editors indicates that significant barriers remain to facilitating critical analysis and debate about evidence syntheses and problematic trials. Our experience of communicating our concerns to the editors of the journals that published the NMAs that included Attari et al. raises important questions about the responsibility of authors and editors of evidence syntheses. If editors have no responsibility to alert readers to such concerns, but the authors of the original trials ignore correspondence, it is not clear what recourse there is (or should be) for researchers when they encounter such findings. At the time of writing we are raising our concerns with the publishers.

### Strengths and Weaknesses

We were unable to replicate all numerical estimates exactly, and we did not have access to all original code and data used in the three NMAs. Discrepancies in our results are minor, however, and are likely to be caused by small variations in statistical modelling (E.g., different methods for node-splitting[28]). For Hetrick, there may have been minor variations in our data extraction compared to the original NMA. However, we were able to reproduce the principal conclusions and inferences, i.e. the different magnitude of the effect estimates across the NMAs, and the existence of inconsistencies between direct and indirect estimates in the Lancet NMAs. These issues were resolved by the exclusion of Attari et al. and this was confirmed in sensitivity analysis using both Bayesian and frequentist approaches.

Several other studies have reported on the effects of problematic trials in evidence syntheses, with sometimes substantial changes in effect sizes or even a change of direction of the effect.[29,30] We are not aware of a systematic review of how the inclusion/exclusion of problematic trials in meta-analyses can alter treatment recommendations, and of how the publishing system corrects (or does not correct) systematic reviews and meta-analyses when problematic trials are detected after publication. However, the negative effects of problematic trials can only be mitigated effectively if authors and editors involved in the affected evidence syntheses take responsibility for making corrections.[31]

### Implications for Practice and Research and Unanswered Questions

Our results may help clinicians and guideline developers who use evidence syntheses to estimate the efficacy of fluoxetine and weigh up the risks and benefits of its use in this population. Our re-analyses give a lower point estimate with narrower certainty intervals that exclude much of the lower portion of the original Lancet NMA credible intervals, and more closely accord with the results of Hetrick. Furthermore, our results bring the estimates for the Lancet NMA datasets (excluding Attari et al.) largely within the range of clinical equivalence as defined by Hetrick.

Our findings emphasise that confidence ratings in risk of bias analyses may not communicate effectively the degree to which the data underlying NMA results may be unreliable. Furthermore, confidence ratings are often only a detail in the results section or supplementary information. It remains an open question how researchers can avoid such important contextual information being lost in evidence summaries and guidelines. Sometimes ‘low confidence’ should be replaced with ‘no confidence’.

Our re-analysis has implications for the broader field of evidence synthesis and medical publishing. It highlights the importance of post-publication peer review and how transparency in the process of analysis, including the sharing of datasets and code, is important to facilitate this process. Our experience of raising concerns with authors and editors highlights the difficulty of ensuring transparency and critical appraisal of the published evidence when concerns about trials and their impact on evidence syntheses arise. While the need for self-correction in the publishing system is acknowledged, open questions remain as to how this process can be enforced and expedited, especially in cases where treatment recommendations - and patient care - are directly affected.

## Supporting information

PRISMA-NMA Checklist

## Data Availability

The data from two of the three network meta-analysis are publicly available, and all data that we used for the re-analysis of the three network meta-analyses are publicly available (links in text).

https://osf.io/jyq5w/

## ADDITIONAL INFORMATION

### Acknowledgments

We thank Cipriani et al. and Zhou et al. for publicly sharing their data. We thank Jack Wilkinson, lead author of the INSPECT-SR template, for sharing the latest version of the template and providing feedback on our assessment of Attari et al. (2006).

### Contributor and guarantor informationt

Conceptualization, drafting, data extraction, data management, and data-cleaning: RL, MP. Statistical analysis: MP, GvV. Reviewing and revising: RL, FN, GvV, MP. Responsible for overall content and guarantor: MP and RL. The corresponding author attests that all listed authors meet authorship criteria and that no others meeting the criteria have been omitted.

### Copyright/licence for publication

The Corresponding Author has the right to grant on behalf of all authors and does grant on behalf of all authors, a worldwide licence to the Publishers and its licensees in perpetuity, in all forms, formats and media (whether known now or created in the future), to i) publish, reproduce, distribute, display and store the Contribution, ii) translate the Contribution into other languages, create adaptations, reprints, include within collections and create summaries, extracts and/or, abstracts of the Contribution, iii) create any other derivative work(s) based on the Contribution, iv) to exploit all subsidiary rights in the Contribution, v) the inclusion of electronic links from the Contribution to third party material where-ever it may be located; and, vi) licence any third party to do any or all of the above.

### Data sharing

All statistical code is available via the <u>OSF</u>. The data for Cipriani is available through the <u>Oxford</u> <u>University Research Archive</u>. The dataset for Zhou et al. is freely available through <u>Mendeley</u>, the version we used for our analysis is publicly available <u>here</u>. The data we used for the re-analysis of Hetrick et al. is publicly available <u>here</u>.

### Transparency statement

The lead author (the manuscript’s guarantor) MP affirms that the manuscript is an honest, accurate, and transparent account of the study being reported; no important aspects of the study have been omitted, and discrepancies from the study as originally planned/registered have been explained.

