## Appendix for "A Re-Appraisal of Three Network Meta-Analyses to Explain the Discrepancy in Findings for the Efficacy of Fluoxetine for the Treatment of Depression in Children and Adolescents"

**Figure S1.** Standardized mean difference (SMD) of Attari et al. 2006, based on the pre-and post measures reported in their Table 2 (in the original article), and an assumed correlation between pre- and post-measures between 0 and 0.99. An SMD of 4.22 (dashed horizontal line) can only be achieved with nearly perfect correlation r > 0.98 (dashed vertical line).

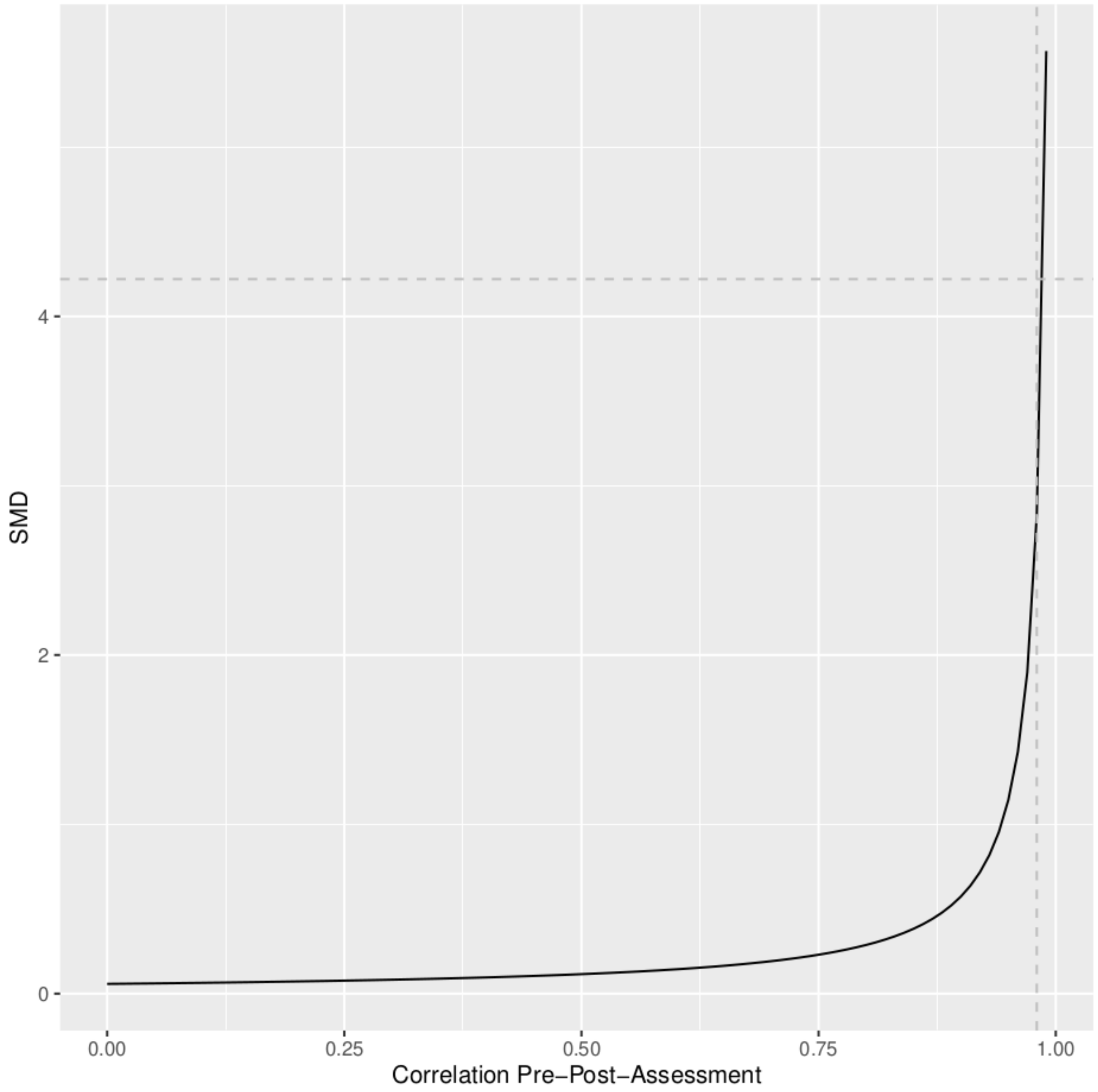

**Figure S2.** Outlier-analyses of Cipriani et al. (2016), Attari et al. (2006) is study #4.

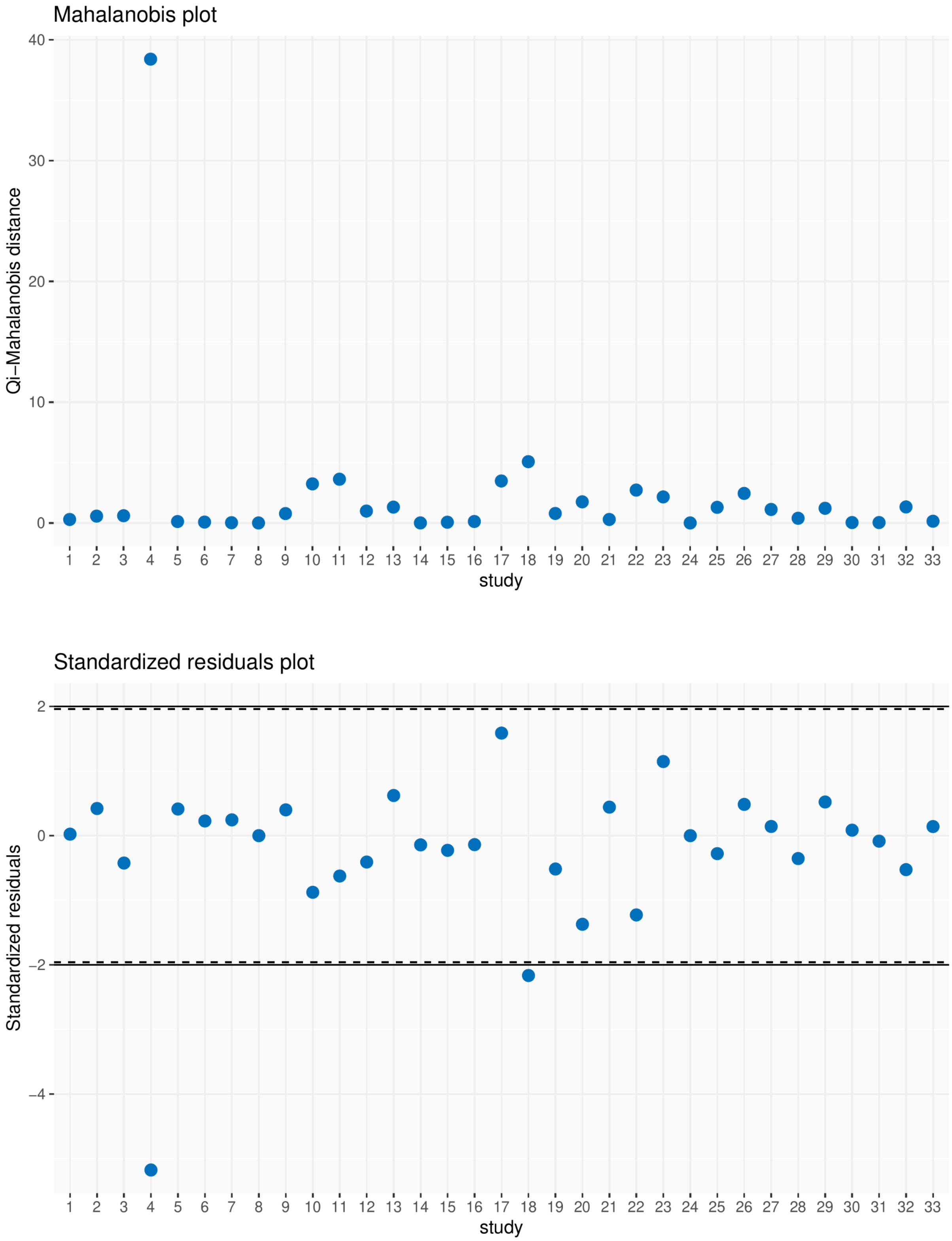

**Figure S3.** Outlier-analyses of Zhou, Attari et al. (2006) is study #13.

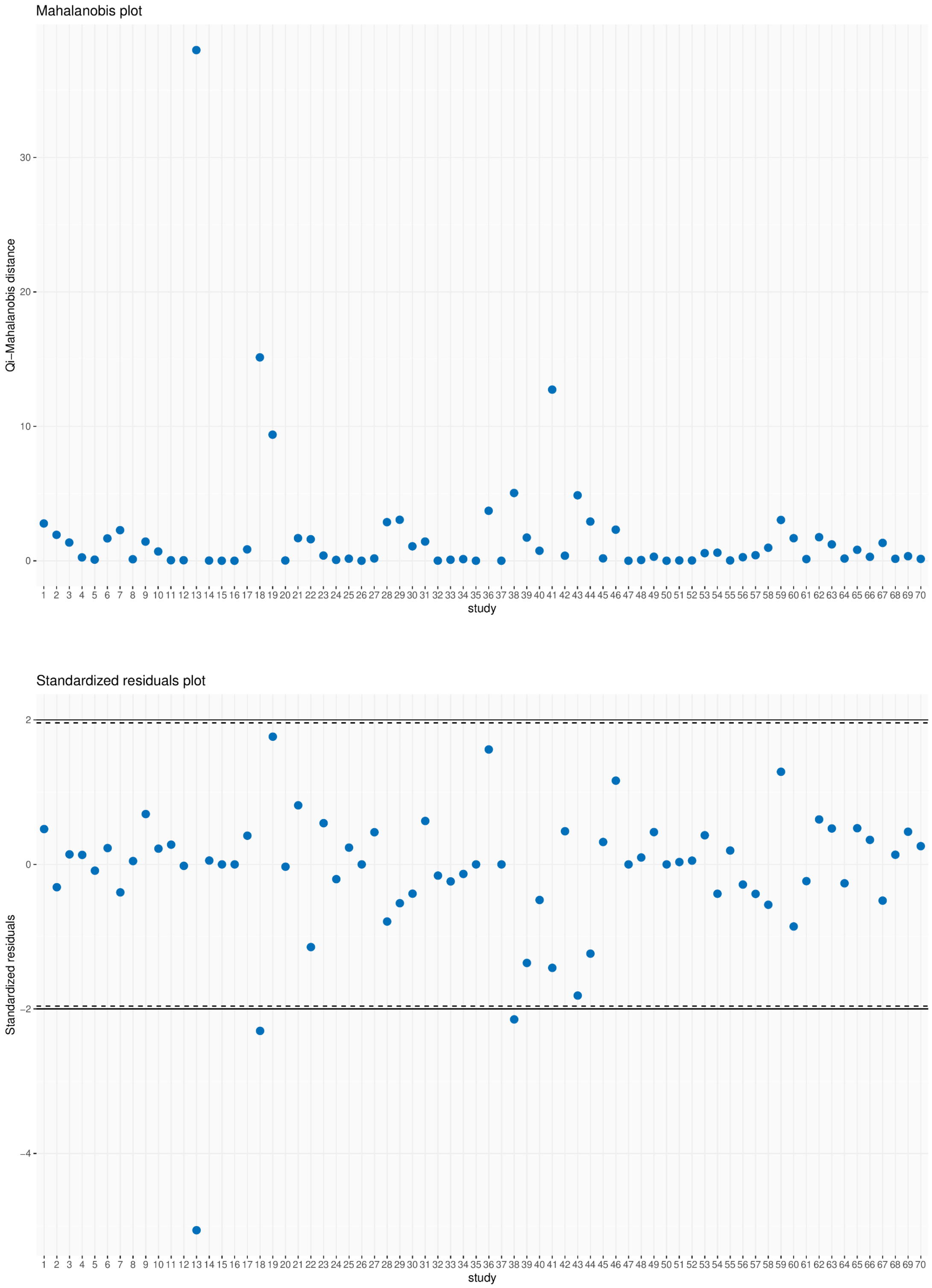

**Table S1.** Results of our sensitivity analyses using a frequentist approach instead of a Bayesian approach for fluoxetine efficacy versus placebo with and without Attari et al. 2006 using the Cipriani et al. and Zhou et al. data.

| **Study** | **Analysis** | **Direct SMD (95% CI)** | **Indirect SMD (95% CI)** | **p-value for the test of incoherence** | **Combined SMD (95% CI)** |
|---|---|---|---|---|---|
| Cipriani 2016 | With Attari | -0.26 (-0.51 to -0.01) | -1.37 (-2.12 to -0.63) | 0.0057 | -0.37 (-0.61 to -0.14) |
| | Without Attari | -0.27 (-0.43 to -0.10) | -0.15 (-0.81 to 0.51) | 0.7162 | -0.26 (-0.43 to -0.10) |
| Zhou 2020 | With Attari | -0.24 (-0.48 to 0.00) | -1.32 (-1.91 to -0.72) | 0.0010 | -0.39 (-0.62 to -0.17) |
| | Without Attari | -0.24 (-0.45 to -0.04) | -0.67 (-1.24 to -0.10) | 0.1669 | -0.29 (-0.49 to -0.10) |
